# Phenotypic Clustering of Systolic Blood Pressure–Heart Rate Synchronization Using Ambulatory Monitoring

**DOI:** 10.64898/2026.01.16.26343820

**Authors:** Maria C Delgado Lelievre, Sabrina Chandra, Ramiro J Valdes Jara, Mehmet Akcin, Carlos L Delgado Leon, Elif Nerez, Elias Hellou, Adam Meyers

## Abstract

**Background:** Interindividual variation in systolic blood pressure (SBP)–heart rate (HR) coupling reflects differences in autonomic and vascular regulation. The primary objective of this study was to determine whether SBP–HR synchronization identifies distinct hemodynamic phenotypes associated with demographic characteristics and blood pressure (BP) profiles in hypertensive and normotensive individuals.

**Methods:** We conducted a cross-sectional cohort analysis of 1,122 adults who underwent continuous 24-hour ambulatory monitoring of SBP, diastolic blood pressure (DBP), and HR in an outpatient clinical setting. Participants were classified as hypertensive or normotensive using guideline-based criteria. The primary exposure variable was SBP–HR synchronization, quantified using zero-lag cross-correlation coefficients. The primary outcomes were demographic characteristics (age, sex, stature) and BP phenotypes, including isolated systolic hypertension (ISH).

Unsupervised partitioning around medoids clustering was used to identify synchronization-based phenotypes. Group comparisons were performed using Welch’s ANOVA and χ² testing, with multivariable adjustment for age and sex where applicable.

**Results:** All 1,122 participants were included in the final analysis (mean age 60.7 years; 52% female). Three synchronization phenotypes were identified: low, moderate, and high. Compared with the low-synchronization phenotype, the high-synchronization phenotype was younger (55.2 ± 15.0 vs. 66.6 ± 13.9 years), taller (170.4 ± 9.4 vs. 167.4 ± 10.5 cm), more frequently male (56% vs. 38%), and had lower baseline SBP (131.8 ± 17.5 vs. 136.7 ± 24.2 mmHg). Overall hypertension prevalence did not differ across phenotypes; however, ISH was less frequent in the high-synchronization phenotype (15.7% vs. 23.4%). Cluster assignments were robust across sensitivity analyses.

**Conclusions:** SBP–HR synchronization identifies distinct hemodynamic phenotypes associated with age, sex, stature, and BP characteristics. Stronger synchronization reflects a physiological profile consistent with preserved autonomic–vascular integration and lower prevalence of isolated systolic hypertension, supporting its potential role in refined cardiovascular phenotyping.

## Introduction

The cardiovascular system functions as an integrated regulatory network in which BP and HR interact continuously to maintain hemodynamic stability.^1–3^ These interactions are mediated by autonomic, vascular, and reflex mechanisms that dynamically adjust cardiac output and vascular tone in response to internal and external stimuli.^4–6^ In hypertension, a condition affecting a substantial proportion of the adult population worldwide^7^, disruption of these regulatory processes contributes to increased cardiovascular risk and end-organ damage.^8–11^ Yet, hypertension is increasingly recognized not as a uniform disorder, but as a collection of heterogeneous physiological phenotypes with distinct regulatory characteristics.^12–15^

The temporal relationship between SBP and HR provides insight into the integrity of autonomic–vascular coupling. Under physiological conditions, coordinated fluctuations in SBP and HR reflect effective baroreflex buffering and balanced sympathetic–parasympathetic control.^16–18^ Conversely, reduced or disordered coupling may indicate impaired autonomic regulation, excessive sympathetic activation, or vascular stiffening.^19–21^ Although circadian BP and HR variability have been linked to adverse cardiovascular outcomes^22–24^, less attention has been directed toward quantifying interindividual differences in SBP–HR synchronization as a phenotypic trait.

Traditional hypertension classification relies primarily on static BP thresholds, which may obscure clinically meaningful differences in underlying hemodynamic regulation.^25^ This limitation has prompted growing interest in data-driven approaches that capture multidimensional physiological patterns rather than isolated measurements. Unsupervised clustering methods, applied to high-resolution cardiovascular data, offer an opportunity to identify subgroups defined by shared regulatory features that may not be apparent using conventional classifications.^26–28^

Despite this progress, the determinants and clinical correlates of SBP–HR synchronization remain incompletely characterized. In particular, the extent to which synchronization patterns vary with age, sex, body stature, and BP phenotype—and whether these patterns associate with specific hypertensive subtypes such as isolated systolic hypertension—has not been systematically examined in large ambulatory cohorts. Clarifying these relationships may improve understanding of cardiovascular regulation across the spectrum of normotension and hypertension.

Accordingly, the primary objective of this study was to determine whether SBP–HR synchronization identifies distinct hemodynamic phenotypes in hypertensive and normotensive individuals. Using continuous 24-hour ambulatory BP and HR recordings, we quantified SBP–HR synchronization and applied unsupervised clustering to define phenotypic subgroups. We then examined associations between these phenotypes and demographic characteristics, anthropometric measures, BP profiles, and hypertension subtypes. We hypothesized that stronger SBP–HR synchronization would reflect a distinct physiological phenotype associated with younger age, male sex, taller stature, and lower prevalence of isolated systolic hypertension.

By integrating dynamic hemodynamic synchronization with phenotypic clustering, this study aims to advance understanding of cardiovascular regulation beyond static BP measurements and to provide a framework for refined hemodynamic phenotyping in clinical and population-based settings.

## Methods

### Study Population

The study cohort consisted of 1,122 adults who underwent 24-hour BP monitoring using FDA-approve non-invasive photoplethysmography-based chest patch monitor^29–31^ in an outpatient clinical setting. Systolic blood pressure, DBP, and HR were recorded at 15-minute intervals. Participants with fewer than 90 valid blood pressure measurements over the monitoring period were excluded to ensure data completeness and quality.

Participants were classified as hypertensive (HT) or normotensive (NT) according to guideline-based criteria. Hypertension was defined as SBP ≥130 mmHg and/or DBP ≥80 mmHg, whereas normotension was defined by SBP <130 mmHg and DBP <80 mmHg. Isolated systolic hypertension was defined by SBP ≥130 mmHg in the presence of DBP <80 mmHg.

### Data Preprocessing and Synchronization Quantification

Time-series data were visually inspected and preprocessed to remove artifacts and ensure temporal alignment between SBP and HR measurements. Missing values were interpolated using linear interpolation when gaps were shorter than 1 hour; longer gaps resulted in exclusion of the affected time segments.

Synchronization between SBP and HR was quantified using zero-lag cross-correlation, equivalent to the Pearson correlation coefficient (r), capturing the instantaneous linear association between the two signals. Positive values indicate concordant SBP–HR fluctuations, whereas negative values reflect inverse coupling.^32–33^

The cross-correlation coefficient was calculated as:

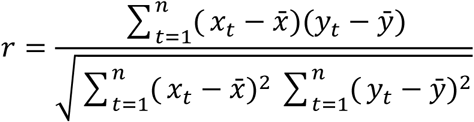

where 𝑥_𝑡_and 𝑦_𝑡_ represent SBP and HR values at time 𝑡, respectively, and *x̄* and *ȳ* denote their 24-hour means.

### Clustering Methodology

Unsupervised clustering was performed using the k-medoids algorithm implemented via the Partitioning Around Medoids (PAM) method.^34^ Clustering was based solely on individual SBP–HR synchronization coefficients (r). This approach was selected for its robustness to outliers and its interpretability, as each cluster is represented by an observed individual (the medoid).

The distance matrix was constructed using the absolute difference between r values. The optimal number of clusters (k) was determined by maximizing the average silhouette width *s̄* across candidate solutions with 𝑘 = 2to 9. The silhouette width for each participant 𝑖was defined as:

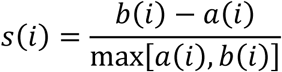

where 𝑎(𝑖)is the mean distance to other members of the same cluster and 𝑏(𝑖)is the minimum mean distance to members of the nearest neighboring cluster. The three-cluster solution yielded the highest average silhouette width and was retained for subsequent analyses.^35^

Clusters were categorized as low-synchronization (−0.49 ≤ r ≤ 0.28), moderate-synchronization (0.28 < r ≤ 0.64), and high-synchronization (0.64 < r ≤ 0.96).

### Covariate Analysis

Demographic and clinical variables included age, height, weight, sex, ethnicity, baseline SBP, and baseline DBP. Continuous variables were compared across clusters using Welch’s analysis of variance, followed by pairwise Welch’s t-tests with Bonferroni correction for multiple comparisons. Categorical variables were analyzed using chi-squared tests, with post-hoc pairwise Z-tests when appropriate.

### Statistical Considerations

All statistical tests were two-sided, with a significance threshold of α = 0.05 after adjustment for multiple comparisons. Analyses were performed using R software (version 4.0.3), employing the *cluster* package for PAM clustering and custom scripts for synchronization analyses.

### Validation and Sensitivity Analyses

Robustness of cluster assignments was evaluated through sensitivity analyses that included alternative interpolation methods for missing data (linear versus spline) and exclusion of participants with more than 10% missing measurements. Cluster structure and membership remained stable across all sensitivity scenarios.^35^

### Independent Data Access and Analysis

One author (the corresponding author) had full access to all data included in this study and takes responsibility for the integrity of the data and the accuracy of the data analysis. The corresponding author confirms that the analyses were conducted according to a prespecified analytic plan, that all relevant data were included, and that appropriate measures were taken to minimize bias and ensure data quality and reproducibility.

## Results

### Cluster Selection and Synchronization Phenotypes

Unsupervised clustering of systolic blood pressure–heart rate (SBP–HR) synchronization coefficients identified an optimal three-cluster solution. The average silhouette score was maximized at *s̄* = 0.584for 𝑘 = 3clusters. Although a two-cluster solution yielded a slightly lower silhouette score (*s̄* = 0.578), the three-cluster configuration was selected because it provided improved separation across the full range of Pearson cross-correlation coefficients (−0.49 to 0.96), avoiding aggregation of heterogeneous synchronization patterns into a single group. For all pairwise comparisons among clusters, a Bonferroni-corrected significance threshold of 0.0167 was applied.

The three synchronization phenotypes were defined as low-synchronization (−0.49 ≤ r ≤ 0.28), moderate-synchronization (0.28 < r ≤ 0.64), and high-synchronization (0.64 < r ≤ 0.96). Of the total cohort, 17.1% of participants were assigned to the low-synchronization cluster, 39.0% to the moderate-synchronization cluster, and 43.9% to the high-synchronization cluster. The distribution of SBP–HR synchronization coefficients and cluster membership is shown in Figure 1.

**Figure 1:**
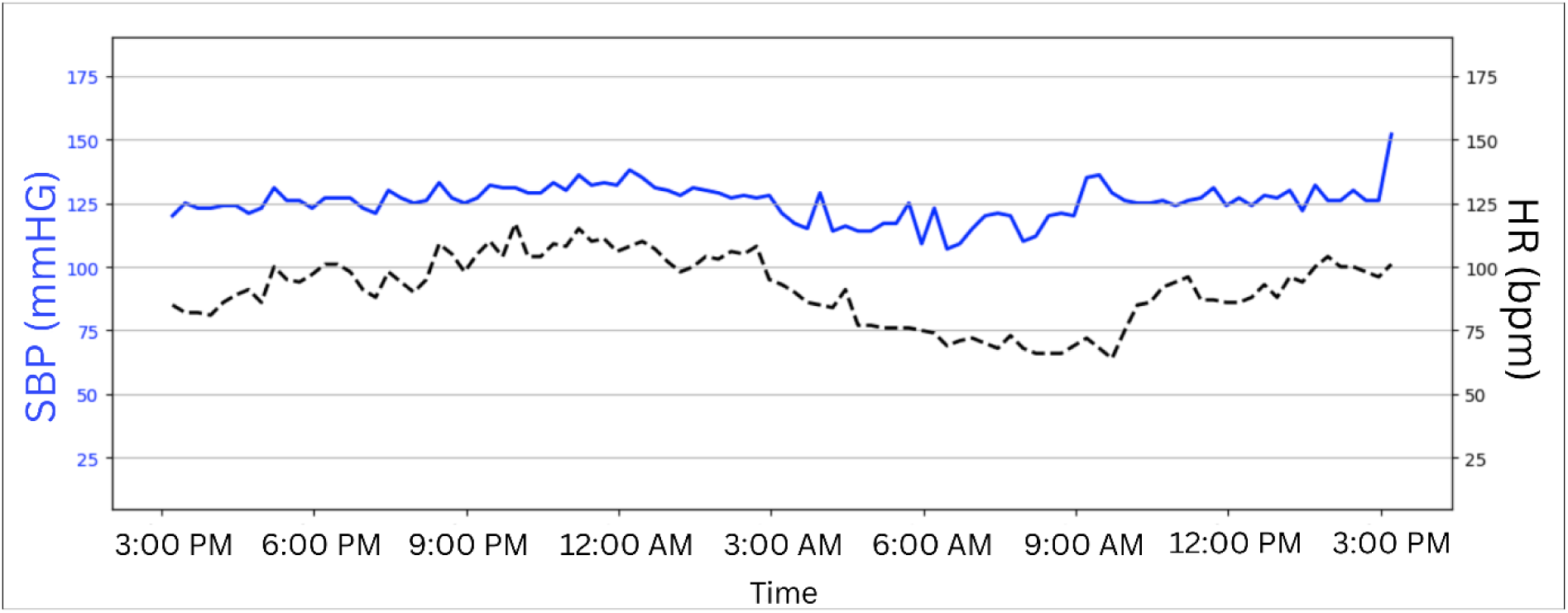
Plot of a sampled patient’s raw (unnormalized) SBP and HR measurements recorded over a 24-hour period. This patient’s readings began at 3:12:55 PM and were sampled every fifteen minutes until the same time the following day.

### Qualitative Synchronization Patterns

Representative SBP and HR time series corresponding to the medoid participant of each synchronization cluster are shown in Figure 2, with signals z-normalized to facilitate comparison of temporal dynamics. In the low-synchronization cluster (Figure 2A), SBP and HR trajectories demonstrated frequent directional discordance over the 24-hour period. Increases in SBP were often accompanied by decreases in HR, and vice versa, consistent with weak or inverse coupling between the two signals. The moderate-synchronization cluster (Figure 2B) exhibited intermediate behavior, characterized by partial directional concordance interspersed with transient dissociations. Periods of coordinated SBP–HR movement were present but not sustained across the full monitoring interval. In contrast, the high-synchronization cluster (Figure 2C) showed closely aligned SBP and HR trajectories throughout the 24-hour period. In this cluster, increases and decreases in SBP were generally accompanied by corresponding changes in HR, indicating strong directional concordance and sustained coupling. Minor amplitude differences and occasional misalignment were observed but did not alter the overall pattern of strong synchronization. Together, these visual patterns illustrate pronounced interindividual variability in SBP–HR synchronization, supporting the existence of distinct hemodynamic phenotypes.

**Figure 2.**
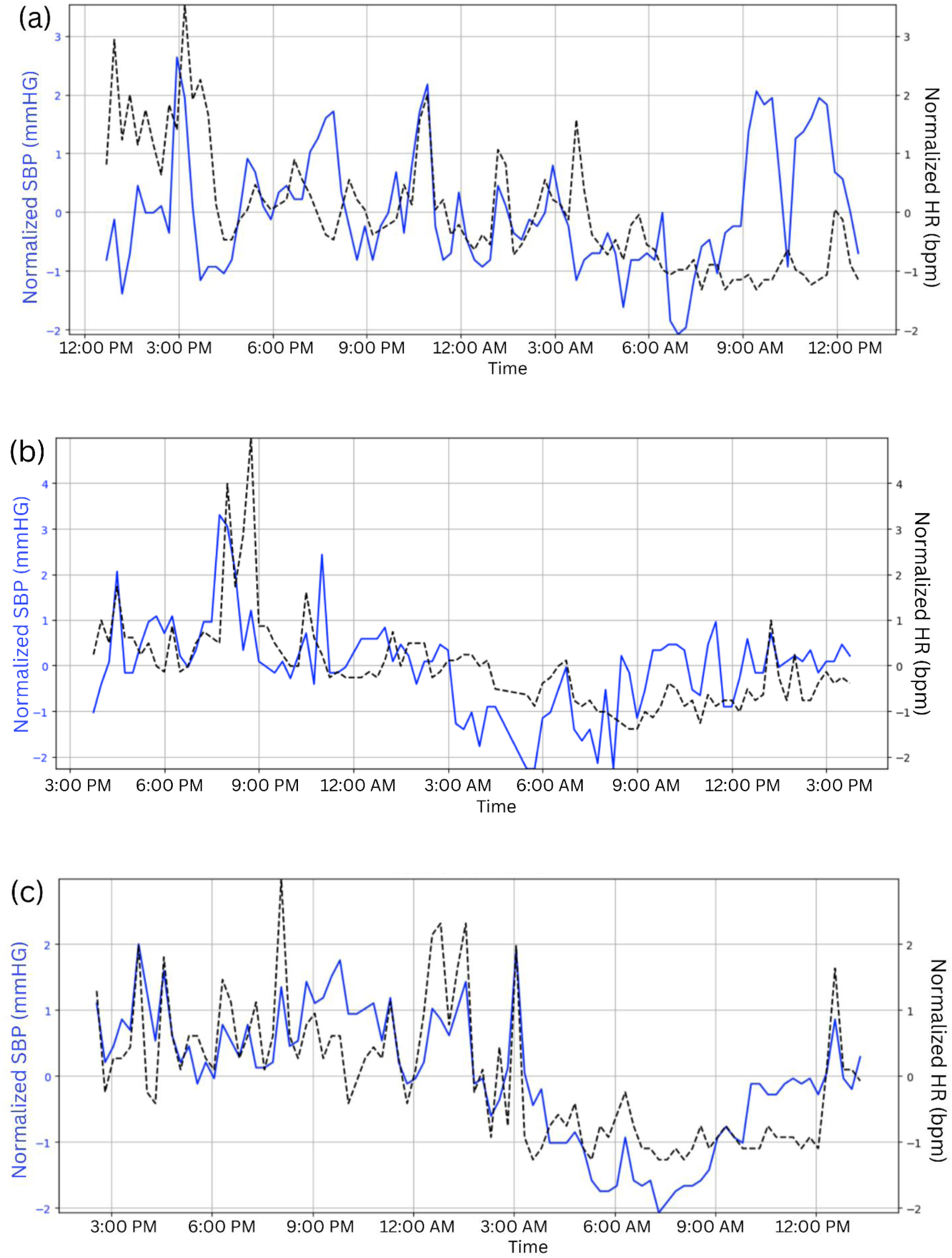
Normalized 24-hour SBP and HR time series corresponding to the patient at the center of the (a) low-synchronization, (b) moderate-synchronization, and (c) high-synchronization clusters, respectively.

### Demographic and Anthropometric Characteristics

Demographic and anthropometric characteristics stratified by synchronization cluster are summarized in Table 1, with results of hypothesis testing presented in Table 2. Mean age differed significantly across all clusters. Participants in the low-synchronization cluster were oldest (66.6 ± 13.9 years), followed by the moderate-synchronization cluster (63.6 ± 14.0 years), and the high-synchronization cluster (55.2 ± 15.0 years). All pairwise comparisons were statistically significant (P*_l-m_* = 0.0113, P*_l-h_* <0.0001, P*_m-h_* <0.0001).

**Table 1.** Summary statistics of clusters.

|  | Cluster<sup>1</sup> |  |  |
| --- | --- | --- | --- |
|  | Low-sync. | Moderate-sync. | High-sync. |
| Range of $r$ values | (-0.49, 0.28) | (0.28, 0.64) | (0.64, 0.96) |
| Patients (% across clusters) | 192 (17.1%) | 438 (39.0%) | 492 (43.9%) |
| Age (years)<sup>2</sup> | 66.6 ± 13.9 | 63.6 ± 14.0 | 55.2 ± 15.0 |
| Height (cm)<sup>2</sup> | 167.4 ± 10.5 | 167.0 ± 10.2 | 170.4 ± 9.4 |
| Weight (kg)<sup>2</sup> | 80.0 ± 17.3 | 80.7 ± 18.3 | 82.0 ± 17.0 |
| Baseline SBP (mmHg) | 136.7 ± 24.2 | 135.3 ± 20.4 | 131.8 ± 17.5 |
| Baseline DBP (mmHg) | 78.1 ± 12.6 | 80.0 ± 11.8 | 80.2 ± 10.5 |
| Sex |  |  |  |
| Male (% within cluster) | 73 (38.0%) | 167 (38.1%) | 274 (55.7%) |
| Female (% within cluster) | 119 (62.0%) | 271 (61.9%) | 218 (44.3%) |
| Ethnicity |  |  |  |
| White (% within cluster)<sup>3</sup> | 30 (22.7%) | 70 (22.7%) | 76 (22.0%) |
| African American (% within cluster)<sup>3</sup> | 10 (7.6%) | 27 (8.7%) | 18 (5.2%) |
| Asian (% within cluster)<sup>3</sup> | 1 (0.8%) | 6 (1.9%) | 3 (0.9%) |
| Hispanic (% within cluster)<sup>3</sup> | 91 (68.9%) | 206 (66.7%) | 249 (72.0%) |
| Hypertensive status |  |  |  |
| Normotensive (% within cluster) | 82 (42.7%) | 199 (45.4%) | 242 (49.2%) |
| Hypertensive (% within cluster) | 110 (57.3%) | 239 (54.6%) | 250 (50.8%) |

<sup>1</sup>Continuous variables are given as mean ± standard deviation. Qualitative variables are given as the raw count and the percentage either across or within clusters.
<sup>2</sup> Means ± standard deviations for Age, Height, and Weight are calculated on 1,094 patients due to the exclusion of outliers.
<sup>3</sup>The percentage of each ethnic group with each cluster is calculated based on the reduced sample sizes of 132, 309, and 346 patients in the low-, moderate-, and high-synchronization clusters, respectively, due to missing data in the variable ethnicity.

**Table 2.** 𝑃-values associated with the global and pairwise hypothesis tests to determine population-level differences between clusters for each covariate.

|  | Global test (*P*)<sup>1</sup> | Pairwise cluster comparison test<sup>2</sup> |  |  |
| --- | --- | --- | --- | --- |
|  |  | Low-Mod.<br>(*P*<sub>*l-m*</sub>) | Low-High<br>(*P*<sub>*l-h*</sub>) | Mod.-High<br>(*P*<sub>*m-h*</sub>) |
| Age (years) | <0.0001* | 0.0113* | <0.0001* | <0.0001* |
| Height (cm) | <0.0001* | 0.6037 | 0.0008* | <0.0001* |
| Weight (kg) | 0.3174 | - | - | - |
| Baseline SBP (mmHg) | 0.0029* | 0.4843 | 0.0100* | 0.0046* |
| Baseline DBP (mmHg) | 0.1201 | - | - | - |
| Sex | <0.0001* | 0.9797 | <0.0001* | <0.0001* |
| Ethnicity |  |  |  |  |
| White<sup>3</sup> | 0.9721 | - | - | - |
| African American<sup>3</sup> | 0.1996 | - | - | - |
| Asian<sup>3</sup> | 0.3994 | - | - | - |
| Hispanic<sup>3</sup> | 0.3377 | - | - | - |
| Hypertensive status | 0.2553 | - | - | - |

<sup>1</sup>Global hypothesis tests use either the ANOVA or chi-squared test and a significance level of 0.05.
<sup>2</sup>Pairwise hypothesis tests include either equality of means or equality of proportions between specified clusters and use a Bonferroni-corrected significance level of 0.0167.
<sup>3</sup>Hypothesis testing for ethnic groups is also performed using the reduced sample sizes of each cluster.
\*Indicates statistical significance.

Height also differed significantly across clusters. Mean height was similar in the low- and moderate-synchronization clusters (167.4 ± 10.5 cm and 167.0 ± 10.2 cm, respectively), whereas participants in the high-synchronization cluster were significantly taller (170.4 ± 9.4 cm). Pairwise testing confirmed significant differences between the high-synchronization cluster and both other clusters (P*_l-h_* =0.0008; P*_m-h_* <0.0001), with no difference between the low- and moderate-synchronization clusters (P*_l-m_* =0.6037). Mean body weight did not differ significantly across clusters (P=0.3174).

### Blood Pressure Profiles

Baseline blood pressure characteristics are presented in Table 1. Mean baseline SBP was highest in the low-synchronization cluster (136.7 ± 24.2 mmHg), intermediate in the moderate-synchronization cluster (135.3 ± 20.4 mmHg), and lowest in the high-synchronization cluster (131.8 ± 17.5 mmHg). Pairwise comparisons confirmed significant differences between the high-synchronization cluster and both other clusters (P*_l-h_* =0.0100; P*_m-h_* =0.0046), while no significant difference was observed between the low- and moderate-synchronization clusters (P*_l-m_* =0.4843).

Mean DBP demonstrated a modest increasing trend with synchronization strength (78.1 ± 12.6, 80.0 ± 11.8, and 80.2 ± 10.5 mmHg in the low-, moderate-, and high-synchronization clusters, respectively), but these differences were not statistically significant at the population level (P=0.1201).

### Sex and Ethnicity

Sex and ethnicity distributions by synchronization cluster are shown in Table 1. The proportion of male participants differed significantly across clusters. The high-synchronization cluster contained a higher proportion of males (56%) compared with the low- and moderate-synchronization clusters (both approximately 38%). Pairwise comparisons confirmed significant differences between the high-synchronization cluster and both other clusters (P*_l-h_* <0.0001; P*_m-h_* <0.0001), whereas no difference was observed between the low- and moderate-synchronization clusters (P*_l-m_* =0.9797).

Ethnic composition did not differ significantly across clusters. Although the high-synchronization cluster contained a slightly higher proportion of Hispanic participants and lower proportions of White and African American participants, global hypothesis tests showed no statistically significant differences across clusters for White, African American, Asian, or Hispanic ethnicity (all P>0.19).

### Hypertension Status and Subtype Distribution

The distribution of normotensive and hypertensive participants across synchronization clusters is shown in Table 3. The proportion of normotensive individuals was highest in the high-synchronization cluster (49.2%) compared with the low- (42.7%) and moderate-synchronization (45.4%) clusters, whereas hypertensive prevalence showed the opposite pattern. These differences did not reach statistical significance (P=0.2553).

**Table 3.** Composition of clusters based on the three-group hypertension classification – normotensive, isolated systolic hypertensive, and (non-ISH) hypertensive.

|  | Cluster |  |  |
| --- | --- | --- | --- |
|  | Low-sync. | Moderate-sync. | High-sync. |
| Hypertensive status |  |  |  |
| Normotensive | 82 (42.7%) | 199 (45.4%) | 242 (49.2%) |
| Isolated systolic hypertensive | 45 (23.4%) | 90 (20.5%) | 77 (15.7%) |
| Hypertensive | 65 (33.9%) | 149 (34.0%) | 173 (35.2%) |

<sup>1</sup>The qualitative variables are given as the raw count and the percentage within clusters.

Further subclassification of hypertensive participants into iISH and non-ISH hypertension is also presented in Table 3. ISH prevalence (Table 4) differed significantly across clusters at the global level (P=0.0343), with higher proportions observed in the low- and moderate-synchronization clusters (23.4% and 20.5%, respectively) compared with the high-synchronization cluster (15.7%). Pairwise comparisons did not meet the Bonferroni-corrected threshold (P*_l-h_* =0.0168; P*_m-h_* =0.0521), although the observed pattern suggests a lower occurrence of ISH among participants with strong SBP–HR synchronization. No significant differences were observed in the proportion of non-ISH hypertensive participants across clusters (P=0.9158).

**Table 4:** 𝑃-values associated with the global and pairwise hypothesis tests to determine population-level differences between clusters regarding the three-group hypertension classification.

|  | Global test<sup>1</sup> | Pairwise cluster comparison test<sup>2</sup> |  |  |
| --- | --- | --- | --- | --- |
|  |  | Low-Moderate | Low-High | Moderate-High |
| Normotensive | 0.2553 | - | - | - |
| Isolated systolic hypertensive | 0.0343* | 0.4159 | 0.0168 | 0.0521 |
| Hypertensive | 0.9158 | - | - | - |

<sup>1</sup>Global hypothesis tests use the chi-squared test and a significance level of 0.05.
<sup>2</sup>Pairwise hypothesis tests include equality of proportions between specified clusters and use a Bonferroni-corrected significance level of 0.0167.
\*Indicates statistical significance.

## Discussion

In this large cohort undergoing 24-hour ambulatory hemodynamic assessment, we identified reproducible patterns of SBP–HR synchronization and demonstrated that these patterns are associated with distinct demographic and clinical characteristics. The principal findings are threefold. First, SBP–HR synchronization exhibits substantial interindividual variability that can be systematically classified into low-, moderate-, and high-synchronization phenotypes using unsupervised clustering. Second, stronger synchronization is associated with younger age, taller stature, male sex, and modestly lower baseline SBP. Third, participants with high synchronization demonstrate a lower prevalence of ISH, although this association did not remain statistically significant after adjustment for multiple comparisons. Together, these findings suggest that SBP–HR synchronization captures physiologically meaningful heterogeneity not reflected by blood pressure thresholds alone.^36–27^

The observed inverse relationship between synchronization strength and age is consistent with progressive alterations in cardiovascular regulation across the lifespan. Aging is accompanied by changes in arterial compliance, baroreflex sensitivity, and autonomic balance, all of which may impair coordinated pressure–rate regulation.^38–40^ Importantly, the age gradient observed across synchronization clusters was continuous rather than categorical, supporting the interpretation that synchronization reflects a spectrum of regulatory integrity rather than discrete pathological states.

The association between stronger synchronization and greater height was independent of body weight and suggests a potential role for anthropometric factors in modulating cardiovascular control.^42–42^ While the mechanistic basis for this relationship cannot be established from the present data, it is plausible that differences in vascular geometry, wave reflection, or baroreceptor loading contribute to more efficient pressure–rate coupling in taller individuals. Notably, this association persisted at the population level and was not explained solely by age or sex.

Sex-based differences were also evident, with males overrepresented in the high-synchronization phenotype. This finding aligns with established sex differences in autonomic regulation and vascular aging trajectories. Although hormonal status and reproductive factors were not assessed, the observed distribution underscores the importance of considering biological sex when interpreting synchronization-based phenotypes and their potential clinical relevance.^43–45^

Baseline SBP was modestly lower in participants with high synchronization, whereas DBP did not differ significantly across clusters. This pattern suggests that synchronization reflects aspects of vascular and autonomic regulation that are not reducible to static blood pressure levels. The trend toward lower ISH prevalence among highly synchronized individuals further supports a link between impaired synchronization and arterial stiffening^46^, although this observation should be interpreted cautiously given the lack of statistically significant pairwise differences after correction.

Qualitative inspection of cluster medoid time series reinforced these findings. High-synchronization individuals demonstrated sustained directional concordance between SBP and HR across the 24-hour cycle, whereas low-synchronization participants exhibited frequent directional discordance. These differences were observed despite identical normalization procedures, indicating that they reflect intrinsic regulatory behavior rather than scaling artifacts. Collectively, these observations suggest that synchronization metrics capture dynamic cardiovascular control properties not accessible through conventional clinic-based measurements.

Several limitations merit consideration. First, the cross-sectional design precludes causal inference regarding whether impaired synchronization precedes or follows the development of hypertension or vascular remodeling. Second, synchronization was quantified using zero-lag cross-correlation, a deliberately conservative and interpretable metric that may not capture nonlinear or time-shifted interactions. Third, the 15-minute sampling interval, although standard for ambulatory monitoring, may overlook faster autonomic fluctuations. Finally, the cohort’s ethnic composition may limit generalizability, particularly for non-Hispanic populations.

Despite these limitations, the present findings have important implications. They demonstrate that SBP–HR synchronization is a reproducible and interpretable phenotype associated with demographic and hemodynamic characteristics relevant to cardiovascular risk. These results support further investigation of synchronization metrics as tools for phenotyping cardiovascular regulation, particularly in relation to aging and isolated systolic hypertension. Longitudinal studies will be essential to determine whether synchronization patterns predict incident hypertension, progression to ISH, or adverse cardiovascular outcomes. Integration with complementary physiological measures may further clarify the mechanisms underlying synchronization loss and its clinical significance.

## Conclusion

In this ambulatory cohort, systolic blood pressure–heart rate synchronization delineated reproducible hemodynamic phenotypes associated with age, sex, stature, and baseline systolic blood pressure. Individuals with stronger synchronization were younger, taller, more frequently male, and exhibited modestly lower systolic pressure, whereas weaker synchronization characterized older participants and those with features consistent with vascular aging. Although differences in hypertensive status did not reach statistical significance after correction, the observed trend toward lower isolated systolic hypertension prevalence among highly synchronized individuals suggests a potential link between impaired pressure–rate coupling and arterial stiffening. Collectively, these findings indicate that SBP–HR synchronization captures dynamic aspects of cardiovascular regulation not reflected by static blood pressure measurements alone. Further longitudinal and mechanistic studies are warranted to determine whether synchronization phenotypes have prognostic value and can inform individualized cardiovascular risk assessment.

## Data Availability

The data that support the findings of this study are available from the corresponding author upon reasonable request, subject to institutional review and data-use agreements.

## Acknowledgments

none

